# Broad decline and subsequent differential re-emergence of respiratory viruses during COVID-19 pandemic response measures, Singapore 2020

**DOI:** 10.1101/2021.03.23.21251968

**Authors:** Wei Yee Wan, Koh Cheng Thoon, Liat Hui Loo, Kian Sing Chan, Lynette L. E. Oon, Adaikalavan Ramasamy, Matthias Maiwald

## Abstract

**Importance:** COVID-19 pandemic control measures affect the prevalence of other respiratory viruses. Effects on some viruses have been described; however, the broader impact and temporal relationship of control measures on virus decline and subsequent re-emergence have not been thoroughly documented. Understanding these phenomena may influence health policies.

**Objective:** To examine the prevalence of unrelated respiratory viruses in relation to population-wide pandemic response measures and phases in 2020 in Singapore.

**Design, Setting, and Participants:** Data from respiratory multiplex PCRs from 3 major hospitals (total 3700 beds) in Singapore were collated. The full dataset consisted of 42,558 test results, 19,898 from 2019 and 22,660 from 2020.

**Main Outcomes and Measures:** Weekly virus prevalence data were mapped onto prevailing pandemic response measures, in order to elucidate temporal relationships and differential virus responses. Pre-pandemic data from 2019 were compared with data from 2020.

**Results:** Early response measures, even before national lockdown, were followed by a dramatic reduction of influenza viruses and a more gradual decline of other respiratory viruses, including respiratory syncytial virus, parainfluenza viruses, endemic coronaviruses and metapneumovirus. Marked decline of enterovirus/rhinovirus and adenovirus, however, was only observed during lockdown. About 13 weeks into phased reopening, enterovirus/rhinovirus re-emerged, followed by adenovirus, the latter mainly in the pediatric population. All other viruses remained at low levels until the end of 2020.

**Conclusions and Relevance:** COVID-19 control measures in Singapore had a significant impact on a broad range of respiratory viruses. Effects of various control measures varied between phases and different viruses. Influenza viruses declined earliest and most dramatically; relaxation of measures was followed by re-emergence of enterovirus/rhinovirus and adenovirus. These patterns are presumably a result of different propensities for contact versus droplet and overall ease of transmission, and different virus reservoirs. Further studies into these phenomena are a matter of public health importance.

**Key Points:** *Question:* What were the effects of COVID-19 pandemic control measures in Singapore on the prevalence of other respiratory viruses?

*Findings:* Viruses responded differently to control measures. Influenza viruses declined rapidly after early control measures and remained near-absent during reopening after lockdown. Enterovirus/rhinovirus and adenovirus declined later and re-emerged earlier than other viruses during phased reopening.

*Meaning:* Population-wide interventions resulted in a broad decline and subsequent differential re-emergence of non-targeted respiratory viruses, corresponding to different patterns of virus response to control measures.

## Introduction

The COVID-19 pandemic and the rapid spread of SARS-CoV-2 brought unprecedented challenges to the world. Many countries and jurisdictions responded by implementing pandemic control measures that included border closures, lockdowns, school and business closures, travel restrictions, mask-wearing mandates and social distancing measures. This led to “collateral” effects in terms of reduction of other respiratory viruses. Effects on influenza viruses have been reported from various countries.^1–8^ The decline of influenza has also been the topic of a systematic review.^9^ Additional reports have documented the decline of several other respiratory viruses.^10–12^ However, the broader impact on other respiratory viruses, as well as the temporal relationships of control measures on virus decline and subsequent re-emergence, the latter during relaxation of measures, have not yet been well documented.

Singapore went through various pandemic control measures which were implemented in phases (Table 1). The first COVID-19 case was reported on January 23, 2020, in a traveler from China; this was followed by more travel-associated and then locally transmitted cases. The initial response included early detection of cases, isolation and contact tracing. On February 7, the national outbreak alert level was raised to Pandemic Level 3 (of 4 levels). This included limits on large gatherings, suspension of extracurricular activities in schools and advice on personal hygiene. International travel restrictions were implemented on March 24. A national lockdown, termed “Circuit Breaker,” was implemented from April 7 to June 1. This included school and business closures and mandatory mask-wearing outside of home. This was followed by a re-opening in phases (Table 1), during which schools and businesses re-opened and restrictions were gradually eased. The Singapore response included a few unique features, such as public “Safe Distancing Ambassadors,” digitalized check in/out at malls and businesses and the use of digital contact tracing (smartphone app and wearable tokens). During reopening, many of the basic preventative measures, including compulsory mask-wearing outside the home and safe distancing, remained in place.

**Table 1.** Timeline of pandemic response and lockdown measures, Singapore 2020^a^

| Week<sup>b</sup> | Phase | Key events or measures |
| --- | --- | --- |
| 4 | NA | • First imported case reported |
| 6 | Pandemic level 3 (DORSCON Orange) | • Non-essential large events (>1,000 people) deferred<br>• Daily health monitoring at workplace (including temperature)<br>• Businesses encouraged to adopt telecommuting<br>• Schools to suspend external activities and large gatherings<br>• Preschools and social services limit number of visitors<br>• Public reminded of personal hygiene and social distancing<br>• Persons who are unwell to seek medical attention and wear masks |
| 13 |  | • Social gatherings limited to 10 persons<br>• Closure of entertainment venues (e.g. cinemas, bars)<br>• International travel restrictions and stay-home notices for returning residents |
| 15 | Lockdown (“Circuit Breaker”) | • F&B outlets: no dine-in, only takeaways/deliveries<br>• Suspension of non-essential businesses<br>• Restricted commute, work from home encouraged<br>• School closures – move to full home-based learning<br>• Full closure of recreational venues (e.g. museums, theme parks)<br>• Re-usable masks distributed free to all households<br>• Use of masks strongly encouraged outside of home<br>• Deployment of public Safe Distancing Ambassadors |
| 16 |  | • Use of masks mandatory outside of home (anyone >2 years old) |
| 17 |  | • Further restriction on businesses, e.g. hairdressers<br>• Temperature screening & contact traceability logs at all outlets<br>• No social home visits/gatherings |
| 23 | Re-opening Phase 1 | • Phased re-opening of selected businesses<br>• Phased re-opening of schools |
| 25-52 | Re-opening Phase 2 | • F&B outlets: dine-in resumed; no more than 5 persons per table<br>• Safe distancing measures remain in place<br>• Masks outside of home remain compulsory |

Abbreviations: NA, not applicable; DORSCON, Disease Outbreak Response System Condition; F&B, food and beverage.
<sup>a</sup> Source: Ministry of Health Singapore and Wikipedia.<sup>13,14</sup>
<sup>b</sup> Calendar (epidemiological) week in 2020.

## Methods

Data from respiratory virus multiplex PCRs performed during 2019 and 2020 were obtained from three public hospitals in Singapore: Hospital A, the only public specialist women’s and children’s hospital (800 beds), Hospital B, a public hospital offering pediatric and adult medical services (1200 beds), and Hospital C, the largest adult public hospital (1700 beds). These three hospitals represent about 40% of all public hospital beds.^15^ Weekly numbers of tests and positive results for each virus were collated; all were de-identified without patient demographics.

Specimens from patients with respiratory symptoms, sent for routine diagnostic purposes, were tested using the FilmArray Respiratory Panels (BioFire, Salt Lake City, UT, USA; Hospital A, entire period; Hospital B, from May 2020), the NxTAG Respiratory Pathogen Panel (Luminex, Austin, TX, USA; Hospital B, until May 2020) and the Anyplex II RV16 Detection kit (Seegene, Seoul, South Korea; Hospital C, entire period).

## Results

The full dataset comprises 42,558 test results from three hospitals (19,898 in 2019 and 22,660 in 2020). During 2019, influenza A/B and enterovirus/rhinovirus were most commonly detected across the three hospitals, followed by respiratory syncytial virus (RSV) (Figure 1, Table 2). Enterovirus/rhinovirus and RSV rates were higher in Hospital A, which is mainly serving a pediatric population, than Hospitals B/C (eTable). There was a noticeable peak of influenza activity during December 2019/January 2020, corresponding to the Northern hemisphere influenza season. Similarly, enterovirus/rhinovirus activity was high since the second half of 2019, and adenovirus activity was higher than average since about September.

**Table 2.**
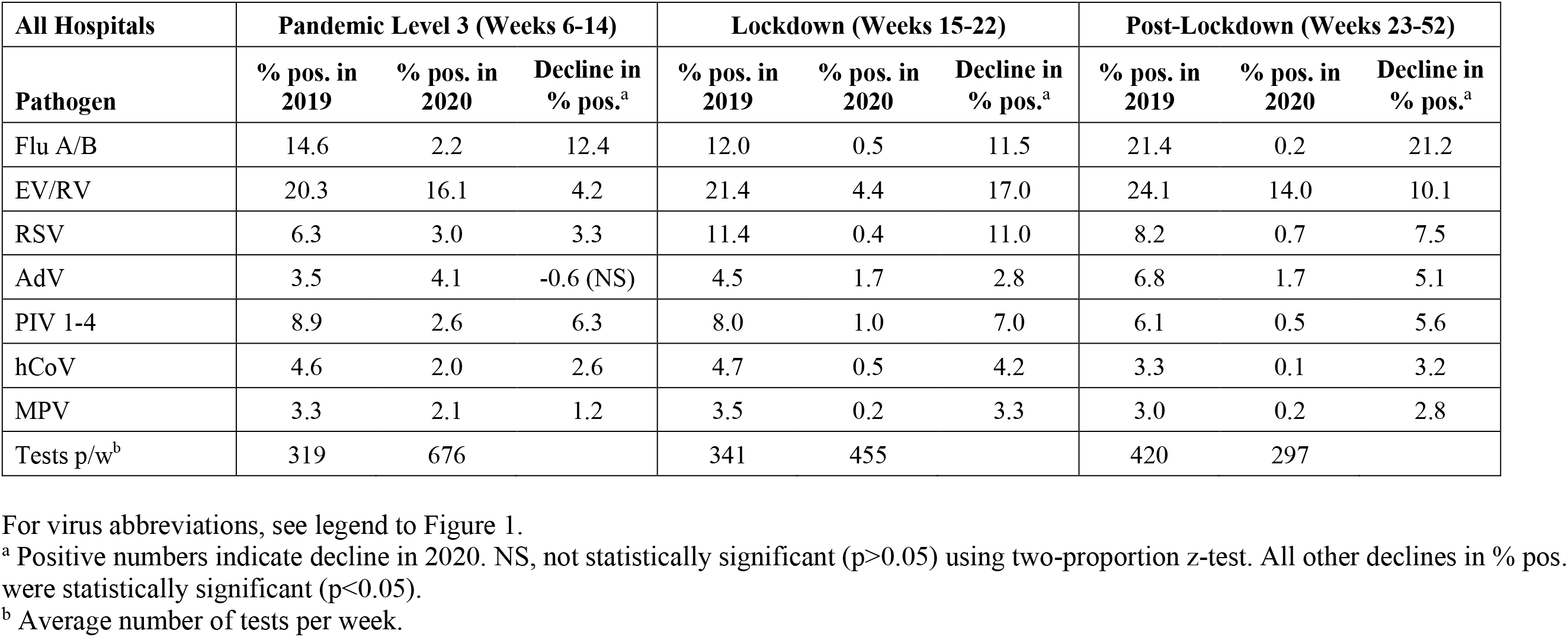
Percentage of positive results for respiratory viruses in weeks 6 to 52 of 2019, and differences in positive results for the same period in 2020, before, during and after the lockdown in Singapore, from all three hospitals combined

| All Hospitals | Pandemic Level 3 (Weeks 6-14) |  |  | Lockdown (Weeks 15-22) |  |  | Post-Lockdown (Weeks 23-52) |  |  |
| --- | --- | --- | --- | --- | --- | --- | --- | --- | --- |
| Pathogen | % pos. in 2019 | % pos. in 2020 | Decline in % pos.<sup>a</sup> | % pos. in 2019 | % pos. in 2020 | Decline in % pos.<sup>a</sup> | % pos. in 2019 | % pos. in 2020 | Decline in % pos.<sup>a</sup> |
| Flu A/B | 14.6 | 2.2 | 12.4 | 12.0 | 0.5 | 11.5 | 21.4 | 0.2 | 21.2 |
| EV/RV | 20.3 | 16.1 | 4.2 | 21.4 | 4.4 | 17.0 | 24.1 | 14.0 | 10.1 |
| RSV | 6.3 | 3.0 | 3.3 | 11.4 | 0.4 | 11.0 | 8.2 | 0.7 | 7.5 |
| AdV | 3.5 | 4.1 | -0.6 (NS) | 4.5 | 1.7 | 2.8 | 6.8 | 1.7 | 5.1 |
| PIV 1-4 | 8.9 | 2.6 | 6.3 | 8.0 | 1.0 | 7.0 | 6.1 | 0.5 | 5.6 |
| hCoV | 4.6 | 2.0 | 2.6 | 4.7 | 0.5 | 4.2 | 3.3 | 0.1 | 3.2 |
| MPV | 3.3 | 2.1 | 1.2 | 3.5 | 0.2 | 3.3 | 3.0 | 0.2 | 2.8 |
| Tests p/w<sup>b</sup> | 319 | 676 |  | 341 | 455 |  | 420 | 297 |  |

For virus abbreviations, see legend to Figure 1.
<sup>a</sup> Positive numbers indicate decline in 2020. NS, not statistically significant ( $p>0.05$ ) using two-proportion z-test. All other declines in % pos. were statistically significant ( $p<0.05$ ).
<sup>b</sup> Average number of tests per week.

**Figure 1.**
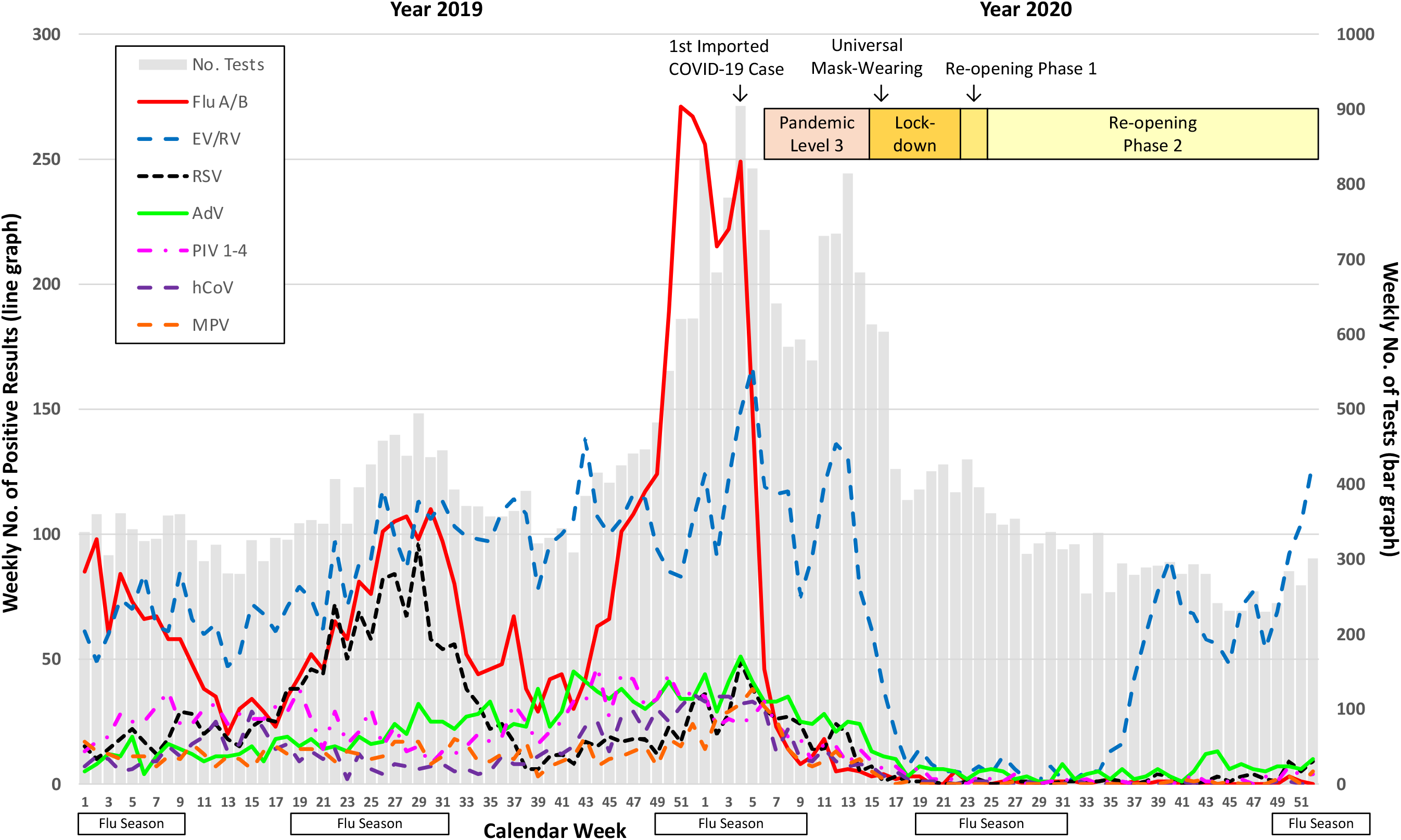
Weekly numbers of respiratory virus multiplex tests performed (grey bars) and of positive tests for different viruses (lines) per calendar week in 2019 and 2020. Influenza seasons are denoted according to Singapore Health Hub.^16^ Abbreviations: Flu A/B, influenza A or B viruses; EV/RV, enterovirus/rhinovirus (combined test); RSV, respiratory syncytial virus; AdV, adenovirus; PIV 1-4, parainfluenza viruses 1-4; hCoV, common cold coronaviruses HKU1, NL63, 229E and OC43; MPV, human metapneumovirus.

Implementation of COVID-19 response measures was followed by marked reductions in detection rates of all respiratory viruses (Figure 1, Figure 2). However, the patterns of decline and subsequent re-emergence differed between groups of viruses and pandemic response phases. Institution of pre-lockdown (Pandemic Level 3) measures was followed by a dramatic reduction of influenza and a more gradual reduction of RSV, parainfluenza, endemic human coronaviruses and metapneumovirus. Marked decline of enterovirus/rhinovirus and adenovirus was only observed after movement restrictions were imposed during the lockdown. When compared to the same time period during 2019, the lockdown phase saw dramatic and statistically significant declines across all respiratory viruses. The Southern Hemisphere influenza season was noticeably absent, there was no mid-year RSV peak as observed in 2019, and there was no Northern Hemisphere influenza season in December 2020.

**Figure 2.**
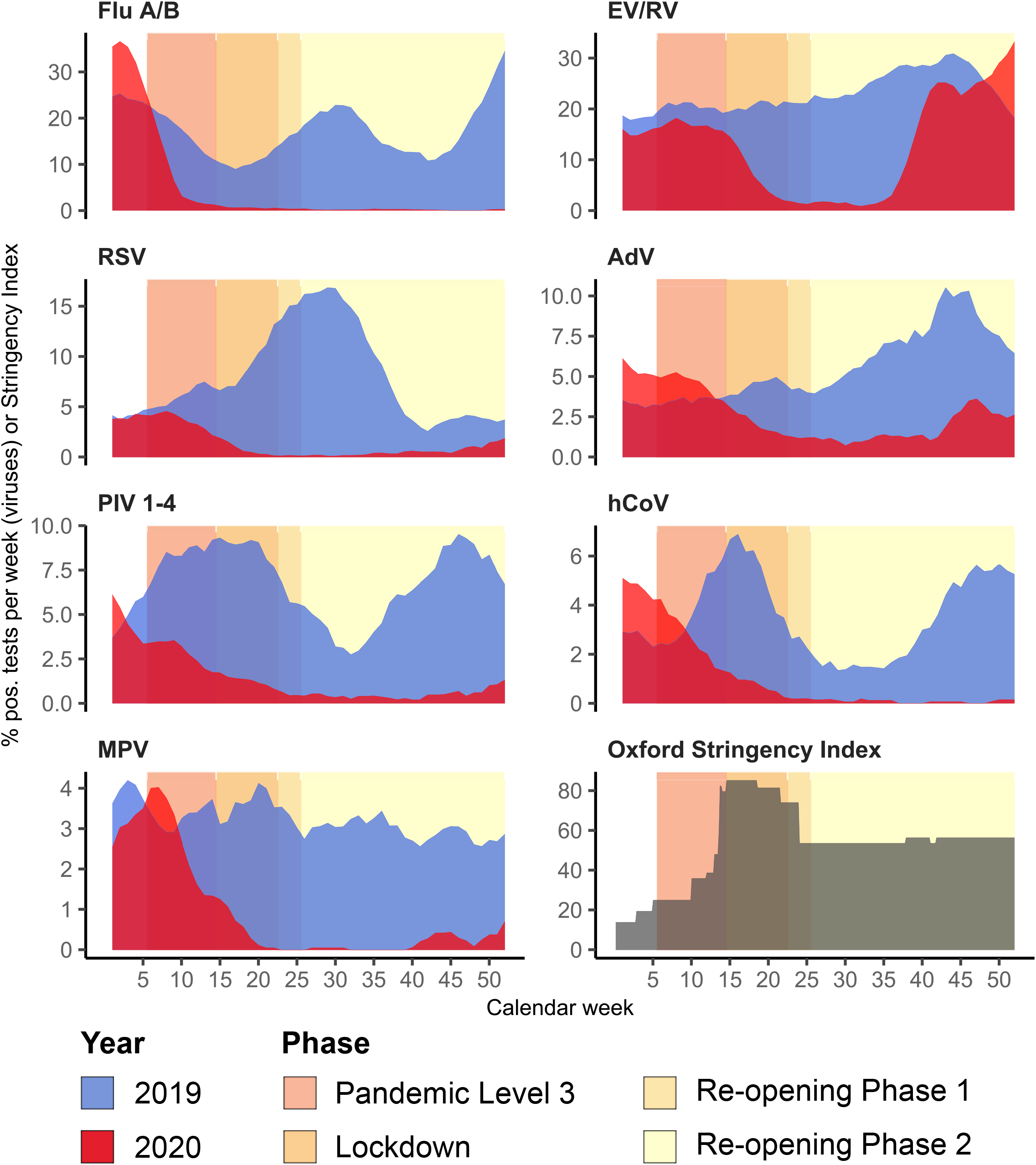
Percentage of tests positive per week for different viruses from three public Singapore hospitals combined for calendar weeks 5 to 52 in 2019 (blue shaded areas) and in 2020 (red shaded areas), plotted as 5-week trailing average, and the Oxford Pandemic Response Stringency Index 2020.^17^ Virus abbreviations as in Figure 1. Percentage scales (y axis) for different viruses vary.

During the reopening phase, low levels of all viruses were sustained for about 13 weeks, but then a marked re-emergence of enterovirus/rhinovirus and a less pronounced rebound of adenovirus occurred in early September and mid-October, respectively. Smaller increases were seen for RSV, parainfluenza and metapneumovirus, but only around November/December. The rebound of enterovirus/rhinovirus and adenovirus was more prominent in Hospitals A/B, which serve a pediatric population, while the adult population (Hospital C) seemed less affected, despite reopening of businesses and gradual resumption of activities (Figure 3). Influenza and endemic human coronaviruses remained at historically low levels until the end of the observation period (December 2020). Generally low levels of influenza, RSV, parainfluenza, endemic coronaviruses and metapneumovirus were sustained despite reopening of schools (with mask-wearing) and the fact that these viruses were common, particularly in children, pre-pandemic.

**Figure 3.**
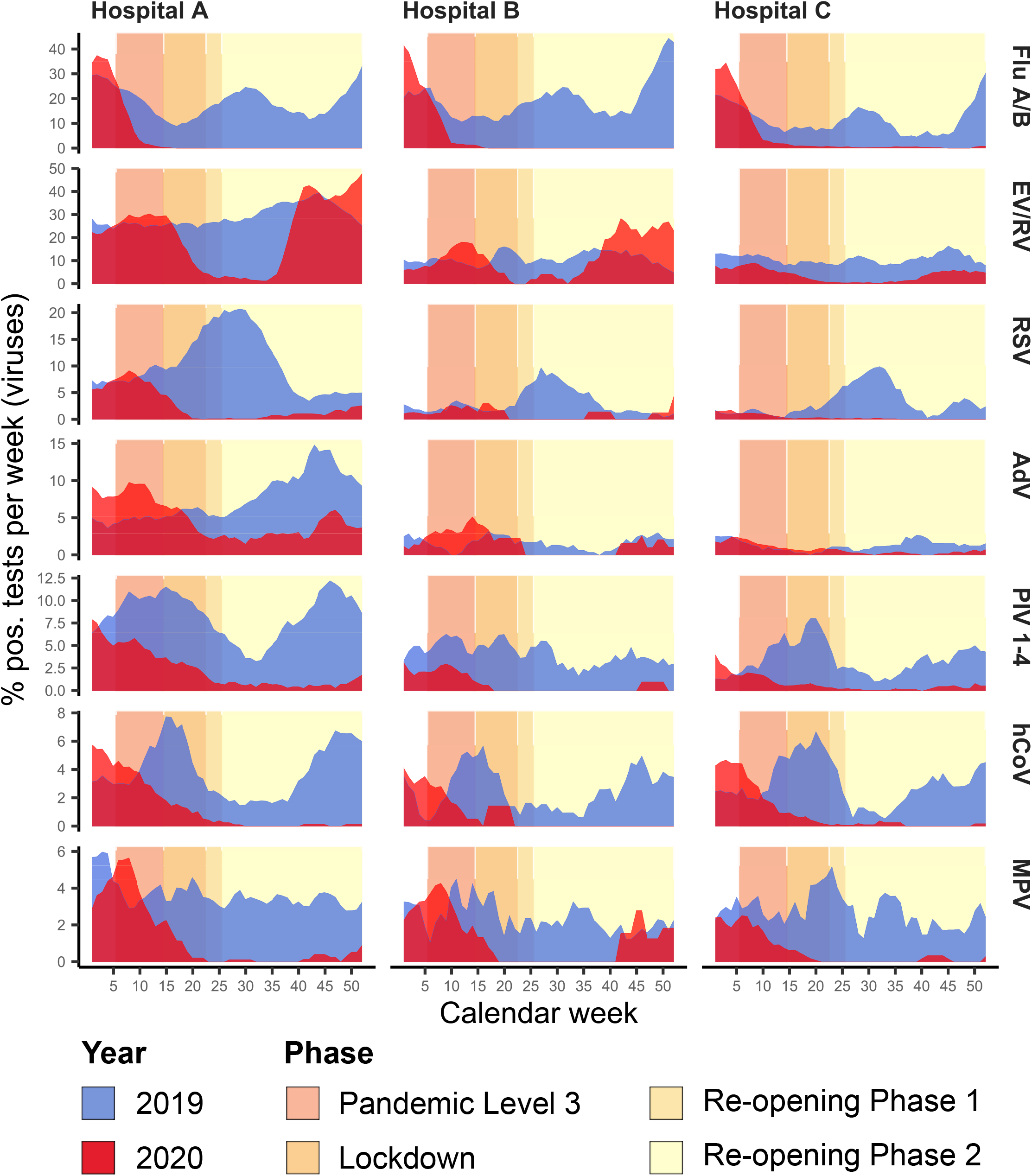
Percentages of tests positive per week for different viruses, shown for individual public hospitals A-C. Virus abbreviations as in Figure 1. Methodology and figure legends as in Figure 2.

Multiplex PCRs in Hospitals A and B did not distinguish between enterovirus and rhinovirus, while the PCR in Hospital C did. A subset of 165 enterovirus/rhinovirus-positive samples from Hospital A from September 2020 was retested using an in-house enterovirus PCR,^18^ and only 16 were positive. Similarly, Hospital C diagnosed 3 enterovirus and 93 rhinovirus infections between June and December 2020. This suggests that the majority of enterovirus/rhinovirus infections during this period were due to rhinovirus.

## Discussion

Here we show that COVID-19 pandemic control measures in Singapore had a dramatic impact on a broad range of respiratory viruses, the extent of which appears to be greater than what has previously been described from other settings. Pandemic control measures affected all tested respiratory viruses; the effects were most dramatic during the lockdown period that included school/business closures, and there were differences between different implementation phases, different viruses and between adults and children. Viruses appeared to have different “susceptibility” to different pandemic control measures, presumably reflecting differences in transmissibility and transmission routes.

Drastic decline of influenza was already seen during Pandemic Level 3, at a time of high influenza activity and following relatively modest control measures, including mask-wearing recommended only for unwell/symptomatic persons. Influenza remained near-absent after the lockdown period, despite reopening of schools/businesses, albeit with continued safe distancing measures and universal mask-wearing. International travel restrictions and reluctance to travel likely contributed to this.

The overall impact of control measures appeared to have been greatest in children (Hospital A; eTable); this included influenza plus common childhood viruses such as RSV and parainfluenza. Among adults (Hospital C), the greatest impact was seen for influenza, enterovirus/rhinovirus and endemic human coronaviruses.

For enterovirus/rhinovirus and adenovirus, substantial reductions were only seen after lockdown, and these viruses rebounded earlier than others. Continued mask-wearing did not appear to prevent this. Both are non-enveloped viruses that have greater stability in the environment, and we hypothesize that greater relative propensity for contact transmission, in addition to droplet transmission, may have contributed to these findings, at a time of increasing social contacts in the re-opening period.^19^ Similar observations have been made in one setting in the UK, where rhinoviruses rebounded following the reopening of schools.^20^ Furthermore, different sizes of remaining virus reservoirs in Singapore’s relatively confined population of about 5.7 million inhabitants may have contributed to this differential re-emergence. The multitude of different enterovirus/rhinovirus and adenovirus types may have facilitated this, with each remaining virus reservoir acting as a “founder population” for resurgence.

This study has several limitations. First, the data were obtained from routine diagnostic tests and were dependent on hospitals’ test requesting patterns. Second, changes in healthcare-seeking behavior during the pandemic may have influenced the trends observed, but the magnitude and direction of these changes cannot be easily ascertained. Third, age information was unavailable, which means that pediatric-adult differences were crudely inferred from the hospitals’ typical patient population.

## Conclusions

In conclusion, Singapore’s COVID-19 response represents a unique situation of effectively enforced population-wide interventions that resulted in a broad decline of non-targeted respiratory viruses. A unique pattern of decline was seen that revealed different sub-patterns of virus transmissibility and intervention responses, presumably depending on different propensity for contact versus droplet, and overall ease of transmission.^19^ With phased relaxation of measures, different viruses again showed different propensities for re-emergence, with most enveloped viruses remaining contained at a time of continued universal mask-wearing. Further studies into these phenomena are a matter of public health importance.

## Data Availability

Original (de-identified) data are available from the authors upon reasonable request.

## Acknowledgments

We thank the Department of Laboratory Medicine, National University Hospital, Singapore, for the generous provision of the respiratory virus data from the hospital.

## Author Contributions

Drs Wan, Ramasamy and Maiwald had full access to all of the data in the study and take responsibility for the integrity of the data and the accuracy of the data analysis. Concept and design: Wan, Thoon, Maiwald. Acquisition, analysis, or interpretation of data: All authors. Drafting of the manuscript: Wan, Maiwald. Critical revision of the manuscript for important intellectual content: All authors. Statistical analysis: Wan, Ramasamy, Maiwald. Administrative, technical, or material support: Wan, Loo, Oon, Maiwald. Supervision: Wan, Thoon, Maiwald.

## Supplementary Online Content

This supplementary material has been provided by the authors to give readers additional information about their work.

**eTable.** Percentage of positive results of individual hospitals A-C for respiratory viruses in weeks 6 to 52 of 2019, and differences in positive results for the same period in 2020, before, during and after the lockdown in Singapore

| Hospital A | Pandemic Level 3 (Weeks 6-14) |  |  | Lockdown (Weeks 15-22) |  |  | Post-Lockdown (Weeks 23-52) |  |  |
| --- | --- | --- | --- | --- | --- | --- | --- | --- | --- |
| Pathogen | % pos. in 2019 | % pos. in 2020 | Decline in % pos.<sup>a</sup> | % pos. in 2019 | % pos. in 2020 | Decline in % pos.<sup>a</sup> | % pos. in 2019 | % pos. in 2020 | Decline in % pos.<sup>a</sup> |
| Flu A/B | 16.6 | 2.0 | 14.6 | 12.6 | 0.0 | 12.6 | 21.7 | 0.0 | 21.7 |
| EV/RV | 25.4 | 29.3 | -3.9 | 26.1 | 10.9 | 15.2 | 31.0 | 25.1 | 5.9 |
| RSV | 8.6 | 6.4 | 2.2 | 15.3 | 0.8 | 14.5 | 10.3 | 1.1 | 9.2 |
| AdV | 4.8 | 8.0 | -3.2 | 6.0 | 4.2 | 1.8 (NS) | 9.5 | 3.2 | 6.3 |
| PIV 1-4 | 10.7 | 4.6 | 6.1 | 9.3 | 2.1 | 7.2 | 7.4 | 0.8 | 6.6 |
| hCoV | 5.2 | 2.9 | 2.3 | 4.6 | 0.9 | 3.7 | 3.7 | 0.1 | 3.6 |
| MPV | 3.5 | 3.1 | 0.4 (NS) | 3.7 | 0.4 | 3.3 | 3.3 | 0.3 | 3.0 |
| Tests p/w<sup>b</sup> | 215 | 262 |  | 237 | 124 |  | 282 | 145 |  |
| Hospital B | Pandemic Level 3 (Weeks 6-14) |  |  | Lockdown (Weeks 15-22) |  |  | Post-Lockdown (Weeks 23-52) |  |  |
| Pathogen | % pos. in 2019 | % pos. in 2020 | Decline in % pos.<sup>a</sup> | % pos. in 2019 | % pos. in 2020 | Decline in % pos.<sup>a</sup> | % pos. in 2019 | % pos. in 2020 | Decline in % pos.<sup>a</sup> |
| Flu A/B | 12.9 | 2.0 | 10.9 | 13.6 | 0.0 | 13.6 | 27.5 | 0.0 | 27.5 |
| EV/RV | 9.0 | 15.5 | -6.5 | 12.1 | 3.4 | 8.7 | 10.1 | 15.8 | -5.6 |
| RSV | 2.2 | 2.2 | 0 (NS) | 2.1 | 1.1 | 1.0 (NS) | 3.7 | 1.0 | 2.7 |
| AdV | 1.0 | 3.8 | -2.8 | 2.1 | 1.1 | 1.0 (NS) | 1.5 | 0.8 | 0.7 (NS) |
| PIV 1-4 | 5.6 | 2.2 | 3.4 | 4.7 | 0.0 | 4.7 (NS) | 3.3 | 0.3 | 3.0 |
| hCoV | 3.9 | 1.4 | 2.5 | 2.7 | 1.1 | 1.6 (NS) | 2.4 | 0.0 | 2.4 |
| MPV | 3.4 | 2.6 | 0.8 (NS) | 2.4 | 0.0 | 2.4 (NS) | 2.1 | 0.8 | 1.3 (NS) |
| Tests p/w<sup>b</sup> | 46 | 72 |  | 42 | 11 |  | 65 | 13 |  |
| Hospital C | Pandemic Level 3 (Weeks 6-14) |  |  | Lockdown (Weeks 15-22) |  |  | Post-Lockdown (Weeks 23-52) |  |  |
| Pathogen | % pos. in 2019 | % pos. in 2020 | Decline in % pos.<sup>a</sup> | % pos. in 2019 | % pos. in 2020 | Decline in % pos.<sup>a</sup> | % pos. in 2019 | % pos. in 2020 | Decline in % pos.<sup>a</sup> |
| Flu A/B | 8.6 | 2.4 | 6.2 | 8.5 | 0.8 | 7.7 | 15.2 | 0.4 | 14.8 |
| EV/RV | 10.3 | 6.2 | 4.1 | 9.7 | 1.9 | 7.8 | 10.3 | 2.3 | 8.0 |
| RSV | 1.0 | 0.7 | 0.3 (NS) | 3.0 | 0.2 | 2.8 | 4.2 | 0.1 | 4.1 |
| AdV | 0.8 | 1.1 | -0.3 (NS) | 0.6 | 0.7 | -0.1 (NS) | 1.4 | 0.3 | 1.1 |
| PIV 1-4 | 4.6 | 1.1 | 3.5 | 5.5 | 0.7 | 4.8 | 3.2 | 0.3 | 2.9 |
| hCoV | 3.2 | 1.5 | 1.7 | 6.1 | 0.4 | 5.7 | 2.6 | 0.1 | 2.5 |
| MPV | 2.5 | 1.2 | 1.3 | 3.6 | 0.2 | 3.4 | 2.4 | 0.1 | 2.3 |
| Tests p/w<sup>b</sup> | 58 | 341 |  | 62 | 320 |  | 73 | 139 |  |

For virus abbreviations, see legend to Figure 1.
<sup>a</sup> Positive numbers indicate decline in 2020. NS, not statistically significant ( $p \geq 0.05$ ) using two-proportion z-test. All other declines in % pos. were statistically significant ( $p < 0.05$ ).
<sup>b</sup> Average number of tests per week.

